# Estimating the risk of hospitalisation and death in England’s remaining unvaccinated population

**DOI:** 10.1101/2022.04.13.22272869

**Authors:** Joseph Shingleton, Steven Dyke, Archie Herrick, Brodie Walker, Thomas Finnie

## Abstract

The roll-out of COVID-19 vaccines in the England has generally been very good – with over 80% of over 12s having received two doses of vaccine by the start of February 2022, and 67% having received a further booster dose. Despite this, there is a small section of the population who remain unvaccinated, either due to lack of access, hesitancy or resistant to vaccination. In this report we estimate that, during 2021, there were approximately 3,500 deaths in unvaccinated people, who could otherwise have reasonably been expected to receive a vaccination. Further, we show that if all of the remaining unvaccinated population in England were to become infected (or reinfected) with the Omicron variant of COVID-19, we would expect to see approximately 11,700 further deaths and 29,600 hospitalisations. These number could fall to 5,300 and 19,600 respectively, if all but the most vaccine resistant individuals become fully vaccinated.

## 1 Introduction

The United Kingdom’s COVID-19 vaccination programme has been largely very successful. Through 2021 and early 2022, over 80% of the country’s adult population received at least two doses of COVID-19 vaccine, and 67% having received a further booster dose [1]. Thanks in part to this extensive vaccine coverage, the UK was able to effectively reduce its COVID-19 incidence hospitalisation ratio and incidence fatality ratio, greatly reducing the burden of COVID-19 on its healthcare system [2].

Despite the extensive and effective vaccine programme, a small but not insignificant population remains hesitant, or even resistant, to receiving a COVID-19-vaccine [3]. This does not include the segment of the population that is unable to access vaccination -potentially arising from the inability to receive and understand information in English, those who are unable to find time to travel to a vaccination clinic and those who are reluctant to engage with institutions of the British state as a result of migration status.

This paper is concerned with the population who are able to access vaccination, but are hesitant. The process in deciding to be vaccinated is complex, and the root causes of hesitancy are varied. Some vaccine hesitant individuals may have had genuine concerns over the safety or efficacy of the vaccine, preferring to wait for more evidence before committing to immunisations. Experiences of healthcare, sociocultural and religious beliefs may also lead to some to decide against vaccination. Others still may have been persuaded into vaccine resistance by misinformation campaigns, designed to discourage individuals from receiving vaccines.

In this paper, we investigate the impact vaccine hesitancy has had, and is likely to continue to have, on the population of England. We will estimate the number of unvaccinated people who died within 60 days of a positive COVID-19 test in 2021, despite having had a reasonable opportunity to have received at least one COVID-19 vaccine. Further, we will estimate the remaining number of deaths in England’s unvaccinated population, under the assumptions that either (a) no further vaccinations are administered, or (b) all but the most vaccine resistant groups received a vaccination.

## 2 Methods

### 2.1 Vaccine hesitant deaths in 2021

Vaccines in England were distributed by age group, with the first vaccines going to the eldest individuals. People who worked in healthcare and those considered ‘clinically extremely vulnerable’, and their carers were also given priority access to vaccinations. In this paper we consider vaccination rates across five age groups: 15-24, 25-44, 45-64, 65-74 and 75+.

Information regarding the dates at which vaccinations were first available for each age group are readily available from NHS and Government press releases. However, the purpose of this work is to estimate the number of deaths occurring in people who had been offered a vaccine and had a reasonable opportunity to have received it. This is likely to be considerably later than the date vaccines first became available, and further, the length of time between first vaccination, and the vaccine being readily available is likely to differ between age groups. To address this, we consider the date at which full vaccination was available to be the date on which 90% of the total second dose vaccines administered by 1^st^ of December 2021 had been given in each age group. We use the National Immunisation Management Service (NIMS) [4] dataset to obtain age stratified vaccination rates.

We consider any death within 60-days of a positive COVID-19 test as a COVID-19 related death. We use UKHSA COVID-19 death data, linked through a unique ID to the NIMS dataset, to identify the number of unvaccinated deaths in each age group. Unfortunately, linkage between the NIMS and COVID-19 deaths datasets is incomplete, leaving a number of deaths with unknown vaccination status. As such, It is likely that our analysis under-estimates the number of vaccinated deaths over the period.

### 2.2 Risk of hospitalisation and death in remaining vaccine hesitant population

The size of the remaining unvaccinated population in England is estimated using the NIMS dataset, along with mid-2021 population Estimates from Office for National Statistics [5]. Estimates from the UKHSA-Cambridge COVID-19 infection nowcasting model are used to estimate the number of unvaccinated individuals in each group who have not had an infection in the last three months, using a process of Iterative Proportional Fitting (IPF) [6].

UKHSA’s COVID-19 vaccine surveillance report, published in week three of 2022 [7], is used to identify the proportion of hospitalisations and deaths occurring in unvaccinated individuals between week 52 of 2021 and week two of 2022. We use the UKHSA-Cambridge COVID-19 transmission model [8] to estimate the total number of infections over the same period. Unfortunately, the age groups in this model do not align with the age groups used in the vaccine surveillance report. As such, we have assumed that there is equivalent risk of hospitalisation and death in similar age groups. These age groups are detailed in table 1.

**Table 1:** Unvaccinated, vaccinated and unlinked deaths after the point where 90% of the 01/12/21 total second dose vaccinations had been administered.

| Age | Date of 90% of total | Vaccinated deaths | Unvaccinated deaths | Unlinked deaths |
| --- | --- | --- | --- | --- |
| 15-24 | 18/10/21 | 5 | 14 | 4 |
| 25-44 | 06/09/21 | 145 | 154 | 94 |
| 45-64 | 30/06/21 | 1,885 | 1,148 | 547 |
| 65-74 | 13/05/21 | 3,085 | 813 | 536 |
| 75+ | 23/04/21 | 11,184 | 1,406 | 1,651 |
| **Total** |  | **16,304** | **3,535** | **2,832** |

We use Bayes’ theorem to estimate the probability of death or hospitalisation given vaccination status:

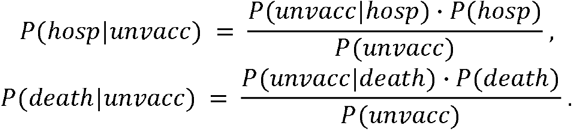

We consider the probability of either outcome given that an unvaccinated individual is infected with COVID-19. We estimate *P*(*unvacc*), the probability of being unvaccinated given you are infected, as the proportion of individuals with any prior infection who have received no doses of vaccine, taken from the results of the IPF analysis.

The probabilities *P*(*hosp*) and *P*(*death*) are taken as the total number of hospitalisations and deaths, divided by the number of infections over the period. We calculate *P*(*unvacc*|*hosp*) and *P*(*unvacc*|*death*) as the ratio of unvaccinated hospitalisations (or deaths) to total hospitalisations (or deaths) in the COVID-19 vaccine surveillance report.

## 3 Results

### 3.1 Estimated vaccine hesitant deaths in 2021

Figure 1 shows the cumulative number of second dose vaccinations in each age group, normalised to 100% of their total value on 1^st^ of December. The vertical lines indicate the date at which 90% of final vaccinations were reached. The calculated dates are given alongside other results in table 1.

**Figure 1.**
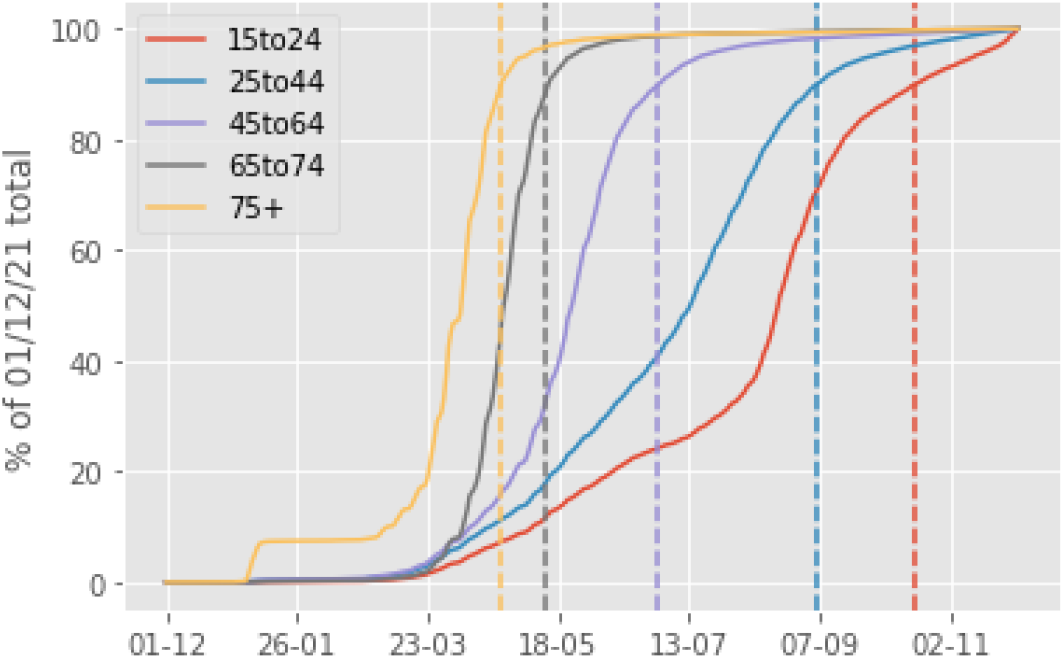
: Proportion of total vaccinations administered on 01/12/21 by date and age group. In the four oldest age groups, vaccinations rate slowed significantly after 90% of the 01/12/21 totals. This was not true for the youngest group, which continued to see growth after this point.

Table 1 shows the number of vaccinated and unvaccinated deaths across each age group, after the point where 90% of total vaccines administered by 1^st^ of December 2021 was achieved. In some cases, it is not possible to link the death and vaccine line lists. These cases are listed as ‘unlinked’ in the table.

The results given in table one show the total number of vaccinated and unvaccinated deaths occurring over the period. However, there are two factors which should be noted here, (a) the time periods are different between age groups, so age comparisons may be difficult, and (b) the vaccinated and unvaccinated population sizes are very different.

These factors have been accounted for in table 2, which shows the number of deaths per day per million people in the unvaccinated and vaccinated population. Since we are looking over the course of several months in some cases, the number of vaccinated individuals is not stationary. We have used the vaccinated and unvaccinated population sizes on 1^st^ of December 2021 as the denominator population, however this choice is arbitrary.

**Table 2:**
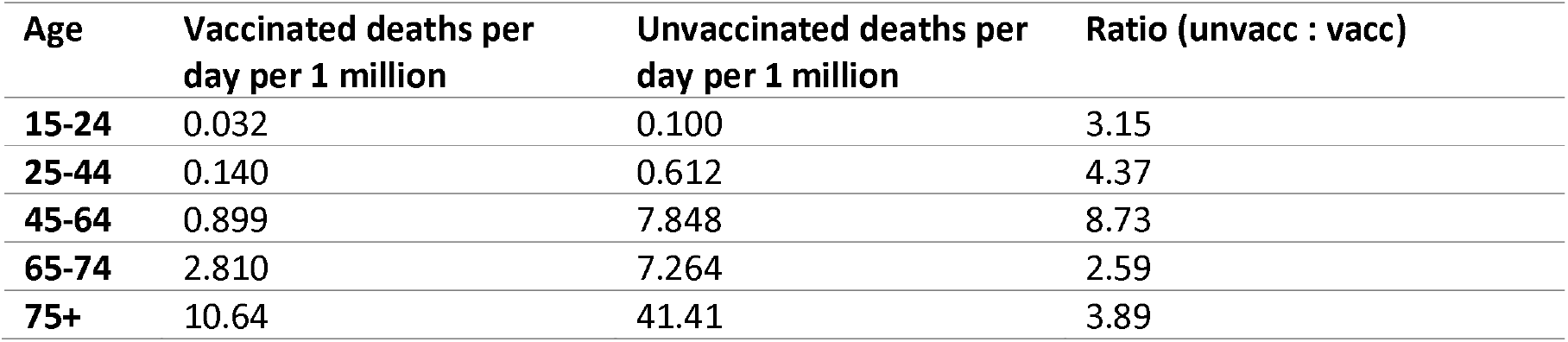
Number of deaths per day per vaccinated and unvaccinated population. Death rates were between 2.6 and 8.7 times higher in the unvaccinated populations.

| Age | Vaccinated deaths per day per 1 million | Unvaccinated deaths per day per 1 million | Ratio (unvacc : vacc) |
| --- | --- | --- | --- |
| 15-24 | 0.032 | 0.100 | 3.15 |
| 25-44 | 0.140 | 0.612 | 4.37 |
| 45-64 | 0.899 | 7.848 | 8.73 |
| 65-74 | 2.810 | 7.264 | 2.59 |
| 75+ | 10.64 | 41.41 | 3.89 |

### 3.2 Estimated risk in remaining unvaccinated population

Table 3 shows the estimated probability of hospitalisation and death, given that an individual is unvaccinated and is infected with COVID-19. Included in this table is an indication of the proportion of the population who are ‘vaccine resistant’ – i.e. they are likely to never receive a vaccine, rather than simply being hesitant or lacking sufficient opportunity to receive a vaccine. These values were taken from an ONS report investigating vaccine hesitancy in the UK [9].

**Table 3:** Probability of outcomes for unvaccinated individuals given infection, independent of testing positive. Note that the estimates for the 75+ age group are inflated by the likely overestimation of vaccine coverage in this group.

| Age group | Real Time Model age group | Percent vaccine resistant | $P(\text{hosp}\vert \text{unvacc})$ | $P(\text{death}\vert \text{unvacc})$ |
| --- | --- | --- | --- | --- |
| 18-29 | 15-24 | 7.9% | 0.003 | 1.2E-04 |
| 30-39 | 25-44 | 6.8% | 0.004 | 3.5E-04 |
| 40-59 | 45-64 | 2.5% | 0.025 | 0.006 |
| 60-69 | 65-75 | 0.5% | 0.103 | 0.049 |
| 70+ | 75+ | 0.5% | 0.081 | 0.058 |

Table 4 shows the estimated number of hospitalisations and deaths in the remaining unvaccinated population, assuming all individuals become infected (or reinfected) with COVID-19. The higher estimate for each age group assumes that no further vaccinations occur in that group. The lower estimate assumes that all but the most vaccine resistant people in each age group receive a vaccine.

**Table 4:** Estimated number of hospitalisations and deaths remaining in the unvaccinated population, given that either no further vaccines are administered (higher estimate) or all but the most vaccine hesitant are vaccinated (lower estimate).

| Age Group | Hospitalisations (low) | Hospitalisations (high) | Deaths (low) | Deaths (high) |
| --- | --- | --- | --- | --- |
| 15-24 | 1,787 | 2,649 | 61 | 90 |
| 25-44 | 3,820 | 4,255 | 356 | 396 |
| 45-64 | 8,912 | 7,901 | 2,017 | 1,788 |
| 65-75 | 3,066 | 4,908 | 1,465 | 2,345 |
| 75+ | 1,980 | 9,929 | 1,413 | 7,086 |
| **Total** | **19,564** | **29,641** | **5,312** | **11,705** |

## 4 Discussion

This study provides an estimate of (a) the number of unvaccinated deaths occurring in 2021 in people we could reasonably expect to have otherwise been vaccinated, and (b) the potential number of deaths and hospitalisations remaining in England’s unvaccinated population. As with any estimation of this kind, our results are based on a number of assumptions.

We use the date at which 90% of total vaccinations by 1^st^ of December 2021 were reached as a reasonable bench-mark for the point at which full vaccination was feasible. In most cases, the rate of vaccine uptake slowed significantly after this point, indicating depletion of the pool of people actively seeking the vaccine. It is at this stage that it is assumed that the remaining unvaccinated are not simply waiting for prescheduled appointments.

However, the decrease in the rate of vaccination was not observed at 90% for the younger age group. This can be attributed to the later availability of the vaccine for the youngest ages (15-17 year olds could only access from the 19^th^ August 2021). This later availability has the opposite effect of the lower uptake of the vaccine in the youngest groups. As such, it is particularly problematic to assume that the number of the unvaccinated deaths were in people who did not have the opportunity to receive a vaccine, rather than due to hesitancy. However, as the relative contribution to total deaths from this group was small we do not expect this choice to have had a significant effect on our findings.

Our results provide estimates for a ‘best’ and ‘worst’ case scenario for the number of unvaccinated deaths remaining in England’s population. Under both scenarios we assume that all individuals in the unvaccinated group will be infected (or re-infected) with COVID-19. While this assumption is reasonable, it may lead to some over-estimation of the risk.

Our analysis has not accounted for re-infections, and the improved prognosis of those infected with partial natural immunity.

Our methods do not account for differences between vaccinated and unvaccinated groups. This is likely to be a concern in the younger age groups, as deaths in these groups will overwhelmingly be in individuals with long term co-morbidities [10]. Vaccine uptake is higher in these groups, given the increased risk of mortality from contracting COVID-19. As a result, the method likely over-estimates the number of hospitalisations and deaths in younger age groups.

Despite the limitations discussed above, our can be expected to provide a reasonable estimate for the number of deaths and hospitalisations remaining in England’s unvaccinated population. By doing this, we provide a degree of understanding of the impact vaccine hesitancy and resistance has on England’s population.

## Data Availability

Some of the data used in this study are not available publicly. Where data are publicly available, links to the relevant sources are included in the bibliography. All data produced by the study are contained within the manuscript.

https://assets.publishing.service.gov.uk/government/uploads/system/uploads/attachment_data/file/1049160/Vaccine-surveillance-report-week-3-2022.pdf

https://www.england.nhs.uk/statistics/statistical-work-areas/covid-19-vaccinations/

